# Varenicline Versus Cytisine for Smoking Cessation in a Primary Care Setting: A Randomized Controlled Trial

**DOI:** 10.1101/2021.07.03.21259600

**Authors:** Stjepan Oreskovic, Jeffrey M. Ashburner, Hrvoje Tiljak, Janez Rifel, Tin Oreskovic, Zalika Klemenc Ketiš, Sanja Percac-Lima

## Abstract

Our study aims to implement a smoking cessation program using pharmacotherapy in a real-life setting — primary care practices in Croatia and Slovenia — and to investigate whether cytisine is at least as feasible and effective as Varenicline in helping smokers to quit in a randomized non-inferiority trial. The use of these medications as an intervention tool for smoking cessation in Croatia and Slovenia is not previously explored and the level of awareness and interest in pharmacotherapy among smokers is unknown.

**Background:** Tobacco addiction is a chronic disorder that accounts for more than 8 million premature deaths each year worldwide^1^. While more than 7 million deaths are the result of direct tobacco use, around 1.2 million are the result of non-smokers being exposed to second-hand smoke.^2^ Smokers suffer from poorer health and are at greater risk of cancer, cardiovascular and respiratory diseases. As a result, half of them die prematurely, on average 14 years earlier.^3^ Among the World Health Organization (WHO) regions, Europe, with 28% of the total population smoking, has the highest prevalence of tobacco smoking in adults and increasing rates among adolescent and female populations.^4^ The European Union and Central Europe in particular has one of the highest proportions of deaths attributable to tobacco use, causing more than 700.000 deaths annually in the European Union.^5^ WHO has estimated that tobacco use is currently responsible for 16% of all deaths in adults over 30 years of age in this region.^6^

Croatia and Slovenia, countries in Central Europe, have a high prevalence of smoking due in part to 1) cultural habits and a tradition of smoking tobacco; 2) problems with financing health care insurance systems; and 3) the strong influence of the tobacco industries on government and national policies. During the past two decades, Croatian and Slovenian tobacco control policies followed the strategies set forth by WHO. However, the impact on the prevalence of smoking differed in each country. Croatia and Slovenia, two European neighboring countries with similar levels of economic development, education and culture have a very high but different smoking prevalence: in Croatia, 39.4% of males and 33.5% of females smoke^7^; in Slovenia, 27.5% of males and 21.1% of females.^8^ Comprehensive tobacco control programs in Slovenia have resulted in significant declines in cigarettes sales and the prevalence of smoking among adults and youth as tobacco control programs have become a national health policy priority.

**Aim:** Our study aims to implement a smoking cessation program using pharmacotherapy in a real life setting — primary care practices in Croatia and Slovenia — and to investigate whether cytisine is at least as feasible and effective as Varenicline in helping smokers to quit in a randomized non-inferiority trial.

**Hypothesis:** Cytisine is at least as feasible and effective as Varenicline in helping smokers to quit in a randomized non-inferiority trial.

**Implications:** The use of these medications as an intervention tool for smoking cessation in Croatia and Slovenia is not previously explored and the level of awareness and interest in pharmacotherapy among smokers is unknown.

Varenicline (trade name: Chantix and Champix) is a prescription medication used to treat nicotine addiction. It reduces craving for smoking cigarettes and other tobacco products. It is a high-affinity partial agonist for the α4β2 nicotinic acetylcholine receptor subtype (nACh) that leads to the release of dopamine in the nucleus acumbens when activated. It has the capacity to reduce the feelings of desire and unpleasant effectsc of the withdrawal caused by smoking cessation^9^.

Cytisine is a generic agent manufactured by Sopharma (trade name Tabex) and by Aflofarm Farmacia Polska (trade name Desmoxan). Cytisine acts by reducing the rewarding effect of nicotine and attenuating nicotine withdrawal symptoms. It’s a plant-based alkaloid found in members of the Leguminosae family (e.g., *Cytisus laburnum*). Cytisine is a partial agonist of nicotinic acetylcholine receptors (nAChRs), binding with high affinity to the α4β2 subtype, which is central to the effect of nicotine on the reward pathway^10^.

Several studies have shown that pharmacotherapy using Varenicline or cytisine is effective for smoking cessation. ^11,12,13^ The current evidence shows that Varenicline is the most effective pharmacotherapy for smoking cessation. Varenicline was more efficacious than bupropion or NRT and as effective as combination NRT for tobacco smoking cessation. It is estimated that Varenicline successfully helps one of every 11 people who smoke remain abstinent from tobacco at six months^14.^ A meta-analysis found that less than 20% of people treated with varenicline remain abstinent from smoking at one year.^15,16^ Four systematic reviews report cytisine to be superior to placebo for short-term and long-term abstinence. The results of the CASCAID trial showed that cytisine was superior to NRT for smoking cessation and, specifically, that cytisine was 1.43 times more likely than nicotine patches with gum or lozenges to help participants stop smoking and remain non-smokers for six months.^11^ However, head to head comparisons in real-life settings are lacking and needed^17^, including a cost-effectiveness analysis taking into the account the fact that cytisine is a low-cost therapy compared with Varenicline and other smoking cessation medications (cytisine at $19.61 for 25 days of treatment; varenicline at $416 for 12 weeks and $832 for 24 weeks). If shown to be non-inferior, therapy with cytisine would be known to result in a lower cost for a similar efficacy.

Primary care practices are an ideal real-life setting for the implementation and evaluation of pharmacotherapy for smoking cessation, as they are exactly the setting where participants would normally receive treatment from their medical care providers, thus addressing possible concerns about generalizations which may limit a study conducted in a context importantly different from that where the investigated treatments would be used.^18^

## Introduction

The European Union and Central Europe, in particular, has one of the highest proportions of deaths attributable to tobacco use. WHO has estimated that tobacco use is currently responsible for 16% of all deaths in adults over 30 years of age in this region^4.^ Croatia and Slovenia, countries in Central Europe, have a high prevalence of smoking due in part to, 1) cultural habits and a tradition of smoking tobacco; 2) problems with financing health care insurance systems, and 3) the strong influence of the tobacco industries on government policies. During the past two decades, Croatian and Slovenian tobacco control policies followed the strategies set forth by WHO; however, the impact on the prevalence of smoking differed between the countries^7,8^ what might be attributable to the gap between the knowledge and current practices^19^ of tobacco control, health promotion and best practices in tobacco related disease prevention programs^21,21^ although numerous research project^22^, interventional studies^23,24^ and internationally recognized health promotion projects^25^ to prevent and control tobacco use and smoking have been implemented in Croatia.

These are the additional reasons why our study aims to assess awareness of and interest in use of pharmacotherapy for smoking cessation and implement a smoking cessation program using pharmacotherapy in a real life setting — primary care practices in Croatia and Slovenia — and to investigate whether cytisine is at least as feasible and effective as Varenicline in helping smokers to quit in a randomized non-inferiority trial.

Prior to starting the treatment, all study subjects, regardless of group assignment, will fill out the Fagerström test for nicotine dependence. Patients will be given medication, explained how it is taken, and a phone number to call with any questions. General medicine doctors, specialized in family medicine and involved in the study are trained in the use of both Varenicline and cytisine, in recognising adverse events and in managing them (eg nausea, insomnia, nausea, and abnormal dreams, constipation, dyspepsia, sleep disorder, vomiting, and flatulence) and in providing behavioural support in a standardised manner. The MDs participating in the study will enter observations including any adverse events at every checkpoint in the study into a log for each participant. The participants will also fill in their observations and record their satisfaction with the treatment at the checkpoints.

The medication treatment with Varenicline will be given by standard medical protocol: we will start Varenicline treatment one week prior to the patient’s target quit date at 0.5 mg once daily and increase it to 0.5 mg twice daily on day 4. On the target quit date (day 8), the dose will be increased to 1 mg twice daily and maintained for 12 weeks ^32.^

Patients assigned to Cytisine will be given a 25 days supply by standard medical protocol and asked to reduce their smoking during the first 4 days of treatment with and aim to quit on the 5^th^ day. We will follow the standard manufacturer’s dosing protocol:^32^ days 1 through 3, one tablet every 2 hours through the waking day (up to six tablets per day); days 4 through 12, one tablet every 2.5 hours (up to five tablets per day); days 13 through 16, one tablet every 3 hours (up to four tablets per day); days 17 through 20, one tablet every 4 to 5 hours (three tablets per day); and days 21 through 25, one tablet every 6 hours (two tablets per day).

During the treatment period, all patients in the study regardless of assignment to Varenicline or cytisine groups will receive weekly calls from a research assistant for the first 5 weeks, then every two weeks for a total of 12 weeks. The RA will inquire about medication adherence and smoking status using the Fagerström test for nicotine dependence, motivate the patient to continue taking medications, encourage them to stop smoking, and ask about possible side effects or other issues. We will hire six RAs for practices in Slovenia and six for practices in Croatia. They will receive clinical research training particularly related to our study, and brief training in smoking cessation counseling. Prior to study start, we will develop a smoking cessation program manual specifying all RA interventions including language to use while talking with patients and how to document the interventions for future analyses.

Patients will be given medication, explained how it is taken, and a phone number to call with any questions. Inclusion criteria for the study: 1) having a primary care physician at one of the 24 primary care practices in Croatia or at one of 19 primary care practices in Slovenia during this study; 2) aged 18 or older; 3) indicate a desire to quit smoking via survey; 4) do not have contraindications to the use of the medicines being tested (according to the summary of product characteristics for Varenicline and cytisine); 5) indicate an interest in pharmacotherapy via survey. Exclusion criteria for the study: 1) persons with cognitive impairment; 2) with a mental disorder, whether treated or not; 3) judged by the field investigators to have insufficient collaboration/adherence in treatment to participate in the trial; 4) considered by field investigators to have frequent adverse reactions to multiple drugs in the previous treatment of acute and chronic diseases; 5) women who are pregnant or breastfeeding.

## Methods

### Study population

The study will be conducted in 48 primary care practices: 27 in Croatia and 21 in Slovenia. Each practice has between 1,275 and 2,375 adult patients.

According to available smoking registries, more than 300 are current smokers in Slovenian and 450 are current smokers in Croatian practices. To enroll 380 participants in our study, we anticipate reaching out to 2100 smokers and asking them to fill the questionnaire. Study research assistants will survey adult patients to assess smoking behavior, and knowledge of and interest in using pharmacotherapy for smoking cessation. Prior to starting treatment, all study participants regardless of group assignment will fill out the Fagerstrom test for nicotine dependence.^26^

### Sample size

The sample for intervention study will comprise of 380 patients/smokers. The primary analyses will use the intention-to-treat perspective, so all smokers enrolled regardless of medication adherence will be analyzed based on the initial group assignment.

### Design Plan

Researchers will randomly assign treatments to study subjects conducting intervention experiment that will include randomized controlled trial.

### Blinding

- Patients will not know the treatment group to which they have been assigned.
- Personnel who interact directly with the study subjects will not be aware of the assigned treatments.
- Personnel who analyse the data collected from the study are not aware of the treatment applied to any given group.

After informed consent is obtained, the patient will be randomized on the spot to the Varenicline or cytisine group. To facilitate the randomization, we will use a random number generator to randomly allocate the Varenicline or cytisine group assignment in a 1:1 ratio within each practice based on the order of enrolment. This pre-specified random allocation by enrollment order will be uploaded to REDCap, so the random assignment will be blinded to the research staff.

### Randomization

Adult patients aged 18 years or older, with a primary care physician in one of the 40 practices, who are current smokers (smoked > 1 cigarette/day during the month before enrolment) who indicated in the survey that they would like to stop smoking and have an interest in pharmacotherapy will be invited to participate in the study. A research assistant will explain study procedures, provide a fact sheet, and obtain informed consent. After informed consent is obtained, the patient will be randomized to the Varenicline or cytisine group. To facilitate the randomization, we will use a random number generator to randomly allocate the Varenicline or cytisine group assignment in a 1:1 ratio stratified by practice based on the order of enrollment. This pre-specified random allocation by enrollment order will be uploaded to REDCap, so the random assignment will be blinded to the research staff.

### Sampling Plan

#### Data collection procedures

This pre-specified random allocation by enrolment order will be uploaded to REDCap, so the random assignment will be blinded to the research staff. The research staff will upload data, also to REDCap, for every patient participating in the survey, filling the questionnaire and the Fagerstrom test.

#### Sample size rationale

The sample size of 380 is what we could reasonably achieve for the study period having in mind the social and behavioural environment created by the COVID-19 pandemic and strict lockdowns in Croatia and Slovenia. In light of this sample size, instead of taking a frequentist approach to non-inferiority testing and pre-specifying a non-inferiority margin, we will adopt a Bayesian perspective in assessing whether the evidence based on the posterior distribution supports the hypothesis that cytisine is at least as effective as Varencline: more details on this are presented in the Analysis plan.

### Variables

#### Manipulated variables

Treatment group assigned (Varenciline or cytisine)

Measured variables:

Age

Sex

Education

Primary Care Practice

Smoking history (number of cigarettes/day)

Fagerstrom Test for Nicotine Dependence (1-2 low dependence; 3-4 low to moderate dependence; 5-7 moderate dependence; 8-10 high dependence.^26^)

Attitude towards quitting smoking, intention to quit:

> from survey responses, one of 10 possible answers to the prompt “Select the statement that best represent your attitude at this moment.”:
>
> 1. “I enjoy smoking and will never think about quitting no matter what happens.”
> 2. “I never think about quitting smoking but perhaps I will someday.”
> 3. “I rarely think about quitting smoking and have no specific plans to quit.”
> 4. “I sometimes think about quitting smoking but have no specific plans to quit.”
> 5. “I often think about quitting smoking but have no specific plans to quit.”
> 6. “I intend to quit within the next 6 months.”
> 7. “I intend to quit within the next 30 days.”
> 8. “I have already started reducing smoking and have decided which day I will quit.”
> 9. “I have already quit smoking, but I am not confident how long I will resist smoking.”
> 10. “I have already quit smoking and I am 100% confident that I will never smoke again.”

Medication adherence at weeks 1, 2, 3, 4, 8, 12, 24 (adherent [>80% of medication taken] or not adherent)

Smoking status at weeks 1, 2, 3, 4, 8, 12, 24 (self-reported 7-day abstinence at each time point)

Continuous smoking cessation at weeks 4, 8, 12, 24

Side effects at weeks 1, 2, 3, 4, 8, 12, 24

### Analysis Plan

#### Statistical models

Feasibility of cytisine treatment will be assessed as % of subjects who are adherent (≥ 80% of medication taken) with their assigned medication at the end of the treatment protocol (cytisine – 4 weeks, Varenicline – 12 weeks) and the estimate and the 95% confidence interval (CI) will be reported. For the smoking cessation outcomes, specifically the comparison at 24 weeks, we will use a Bayesian logistic regression with a single binary predictor indicating the assigned treatment group.

Given the previous literature considering the efficacy of each treatment separately^17^, but a lack of previous direct comparisons, we will choose a recommended^27^ weakly informative prior with the expectation of an estimate close to 0 but some chance of a larger coefficient; a student t distribution with 7 degrees of freedom and a scale of 2.5 will be used for the parameter of interest (the treatment group indicator coefficient). For the primary analysis, we will consider the proportion of the posterior distribution (of the coefficient) that is above or below 0 as the estimated probability that cytisine is at least as effective as Varenicline (or less effective than Varenciline, respectively^28^. Secondarily, we will exponentiate median of the posterior distribution to obtain the point estimate of the odds ratio of the efficacy of cytisine compared to that of Varenicline.

Multivariable logistic regressions will be used to adjust for the Fagerström Test/Score for Nicotine Dependence, age and sex; the multilevel variant of the logistic regression to take into account the country and practice, as well as the repeated measures, in light of the longitudinal data structure. Chi-squared tests will be used for comparisons of other categorical outcomes (e.g. medication adherence, side effects, and interest in pharmacotherapy) between the two groups.

### Transformations

We do not plan any transformations.

### Inference criteria

Statistical significance will be defined as a two-tailed p-value <0.05.

### Data exclusion

Following the original study design, we will exclude data on any participants who will not complete the survey or who are not smokers. Since an intention to treat perspective will be adopted to avoid potential issues arising from non-random attrition, the outcomes will be compared between the participants assigned to the two treatments, regardless of whether the participants, for instance:

- experienced serious side effects
- arbitrarily wished to quit the trial regardless of the reason they cite
- did not adhere to the test protocol

However, since attrition due to becoming pregnant is in fact random, participants who become pregnant during the study will be excluded.

### Missing data

Patients with missing data on smoking status will be assumed to still be smoking.

### Exploratory analysis

We will conduct exploratory secondary analyses to examine possible heterogeneity with regards to age, sex and level of nicotine dependence, which will be included as independent variables in multivariable regression models, while variation across country and practice will be considered with a multilevel model.

## Discussion

In a context of patients’ “normal” health behavior patterns, the primary care practices would be an ideal real-life setting for the implementation and evaluation of pharmacotherapy for smoking cessation. They are the setting where participants would normally receive treatment from their medical care providers. However, providing smoking cessation therapy with cytisine and Varenicline to primary care patients in the time of the COVID-19 pandemic was a particular challenge. Almost all GP specialists participating in the study noticed, observed and came to the conclusion that COVID-19 pandemic has made an important impact on behavior of patients and primary care physicians that, although significant, is hard to quantify and measure.

The possible positive motivation of the study participants to stop smoking might be related to the potential heightened awareness that tobacco use may increase the risk of suffering from serious symptoms due to COVID-19. Early research indicated that, compared to non-smokers, having a history of smoking may substantially increase the chance of adverse health outcomes for COVID-19 patients, including being admitted to intensive care, requiring mechanical ventilation and suffering severe health consequences ^29, 30^. Studies and evidence from China also show that people who have cardiovascular and respiratory conditions caused by tobacco use are at higher risk of developing severe COVID-19 symptoms. ^31^

The possible negative effect might be connected to the fact that during the critical period of the study (September-December 2020 and March-May 2021), after recruitment was completed and the Fagestrom test administered, patients and primary care physicians in both Slovenia and Croatia were living under “lockdown” while facing the threat reflected in two of the globally fastest growing numbers of confirmed COVID-19-related deaths per million (Figure 1). The physicians were occupied with COVID-19 patients and patients were living under stress, and were reluctant to take a risk of leaving their homes and visiting GP’s offices.

**Figure 1.**
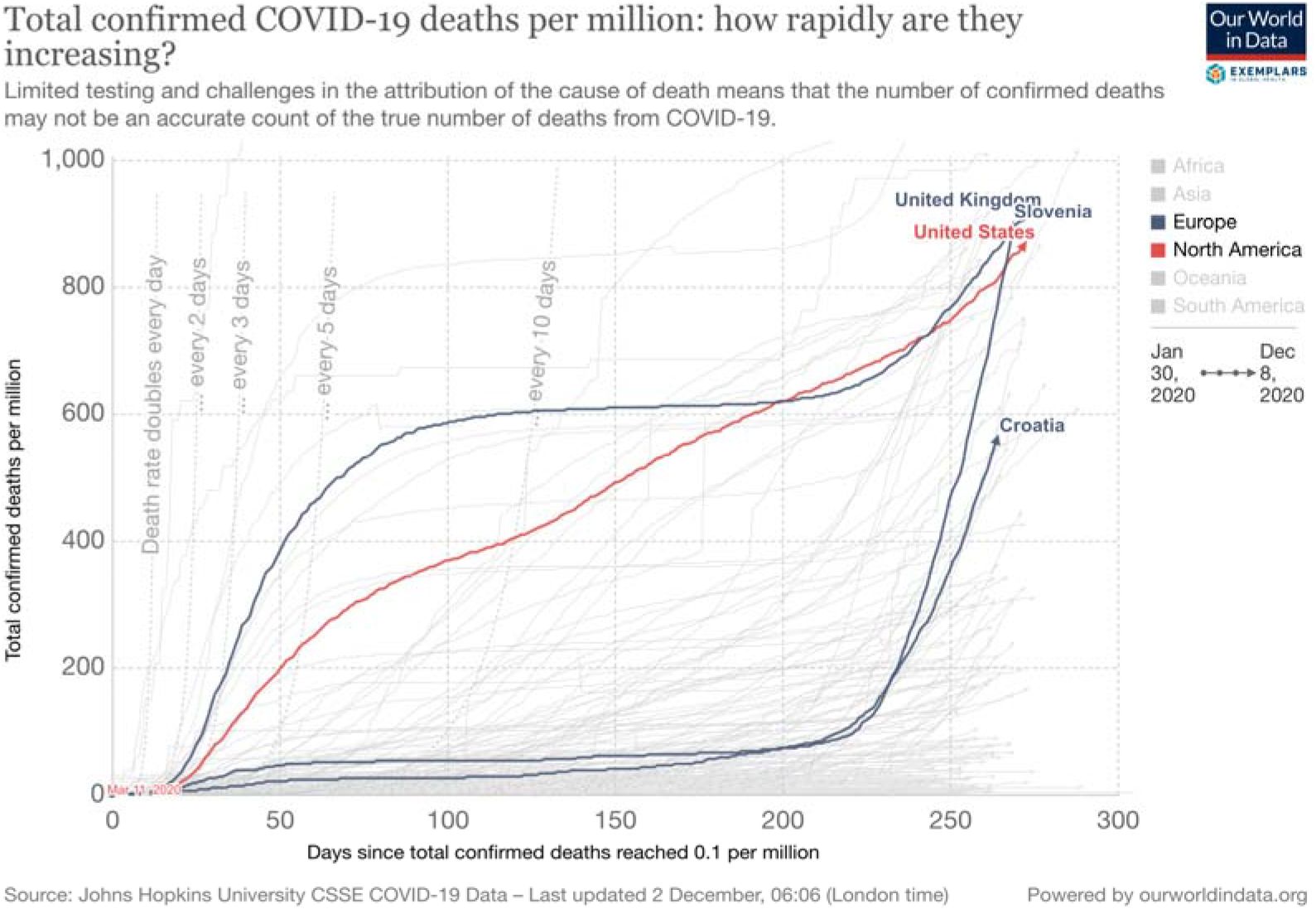
Growth of COVID-19 deaths per million in Croatia and Slovenia in the period of the clinical trial (August-December) as compared to the same period in the USA and the UK

Recruitment started on 14 July 2020, with results expected to be available in September 2021.

## Data Availability

This pre-specified random allocation by enrolment order will be uploaded to REDCap, so the random assignment will be blinded to the research staff. The research staff will upload data, also to REDCap, for every patient participating in the survey, filling the questionnaire, and the Fagerstrom test. 

## Clinical Trial Registration

Trial Registration number: KL:530-07/18-01/72 UB: 381-15/60-18-04

## Study limitations

Smoking status as documented in the practices’ registries may not be up to date, so the number of current smokers could be different than stated in our proposal. We used conservative estimates and believe that we can achieve our recruitment goal. However, we will closely monitor recruitment per practice/country and if lower than anticipated, we will add one practice in Slovenia and one in Croatia to achieve our recruitment target of 380 patients interested in pharmacotherapy for smoking cessation.

## Smoking status will be self-reported

The family medicine practices participating in this study have first class clinicians and staff. They are located in large cities and affiliated with academic medical centers so results might not be generalizable to community/rural family medicine practices.

## Ethical considerations

Trial participants are not seen and receive no reimbursement for their time (although trial medication is free). A two-step verbal consent process is used. Ethics approval was obtained on 13 December 2018 from the National Ethical Committee of the Republic of Croatia. Agency for Medical Products and Medical Devices of Republica of Slovenia ethics approval was obtained on 16 Octobber 2019 following the guidelines of the European Medical Agency and Committee for proprietary Medicinal Products: “Note for Guidance On Good Clinical Practice” CPMP/ICH/135/95)

